# Fed-GLMM: A Privacy-Preserving and Computation-Efficient Federated Algorithm for Generalized Linear Mixed Models to Analyze Correlated Electronic Health Records Data

**DOI:** 10.1101/2022.03.07.22271469

**Authors:** Zhiyu Yan, Kori S. Zachrison, Lee H. Schwamm, Juan J. Estrada, Rui Duan

## Abstract

Large collaborative research networks provide opportunities to jointly analyze multicenter electronic health record (EHR) data, which can improve the sample size, diversity of the study population, and generalizability of the results. However, there are challenges to analyzing multicenter EHR data including privacy protection, large-scale computation, heterogeneity across sites, and correlated observations. In this paper, we propose a federated algorithm for generalized linear mixed models (Fed-GLMM), which can flexibly model multicenter longitudinal or correlated data while accounting for site-level heterogeneity. Fed-GLMM can be applied to both federated and centralized research networks to enable privacy-preserving data integration and improve computational efficiency. By communicating only a limited amount of summary statistics, Fed-GLMM can achieve nearly identical results as the gold-standard method where the GLMM is directly fitted on the pooled dataset. We demonstrate the performance of Fed-GLMM in both numerical experiments and an application to longitudinal EHR data from multiple healthcare facilities.

## 1 Introduction

Electronic health records (EHR) data are valuable for generating real-world evidence in biomedical and epidemiological research [1]. With the increasing availability of EHR data among healthcare facilities [2], integrating these data from multiple institutions has great potential for improving statistical power and generalizability of results [3]. Such integration is also particularly valuable - and often necessary - for studying rare conditions and underrepresented subpopulations [4]. As a consequence, an increasing number of large clinical research networks have been built domestically and internationally to facilitate multicenter EHR-based studies. For example, the Patient-Centered Outcomes Research Institute has launched PCORnet to support a national research collaborative that empowers large-scale comparative effectiveness research [5]. More recently, large collaborative consortia dedicated to investigating clinical and epidemiological questions about COVID-19 have also been formed [6, 7], as timely observational studies based on large integrated EHR data have been increasingly critical for clinical and health policy decision-making in various areas such as treatment evaluation, diagnostic support and healthcare resource prioritization [8–10].

Despite the importance of multicenter EHR-based studies, challenges exist in terms of how to effectively and efficiently compile and analyze multiple large-scale EHR datasets [11]. To overcome data sharing constraints due to privacy regulations and computational constraints, several data-sharing models are commonly used among multicenter research networks. Depending on whether the individual-level data are shared, a research network can be categorized them into either a *federated* or *centralized* network. A *federated* network keeps patient-level data within each institution, and only allows summary-level statistics to be shared across institutions. Some federated research networks allow automated queries, analysis and sharing summary statistics through application programming interfaces and cloud computing, which saves human labor from cross-institutional communication but may require additional safeguard of data breaches [12, 13]. Other federated networks rely on manually transferring summary statistics, which has less requirement on the infrastructure, and is considered more reliable for privacy protection. Thus, this practice is widely adopted among international research networks [6, 14, 15]. In federated networks, federated algorithms are needed to conduct joint analyses across multiple datasets without sharing patient-level data. In contrast, *centralized* networks managed to directly pool deidentified patient-level data across intuitions and store them in centralized data warehouses [16, 17]. When all data are pooled together, fitting a model to a pooled dataset (referred to as the *pooled analysis* hereafter) is feasible but may be subject to challenges from computational complexity and memory bottlenecks due to the large size of the pooled dataset. Therefore, in a centralized network, distributed algorithms are also needed to overcome computational challenges [18, 19].

Most of the existing distributed or federated learning algorithms focus on regression models with independent observations, including logistic regression and Cox regression [20–26]. However, EHR data are longitudinal and correlated in nature, where multiple medical encounters may be associated with the same patient, physician or facility. Therefore, in EHR-based analyses, methods to account for such multi-level longitudinal and correlated observations are necessary. Among many existing methods to address the multi-level correlated data structure, the generalized mixed model (GLMM) is one of the most widely applied methods with great flexibility [27, 28]. To fit GLMM federatively, one straightforward way is to fit separated models locally across sites and aggregate the local estimates through a meta-analysis [29]. Although meta-analysis is straightforward to implement in practice, it has been shown that its accuracy may be suboptimal, especially when rare conditions are included in the model [23]. More recently, a few methods have been developed which consider using site-level random effects to account for heterogeneity across sites. For example, Luo et al. proposed a lossless algorithm for the linear mixed model [30], and a few methods have been proposed for GLMM [31–33]. However, these approaches only consider site-level random effects, which cannot handle repeated and correlated measures within each site. Methods are needed that allow flexible specification of random effects to account for longitudinal or correlated observations at lower levels.

In this paper, we propose an accurate and fast federated algorithm to fit GLMM (Fed-GLMM) with correlated data structures. Our method can be implemented in both federated and centralized networks with different data-sharing constraints. Specifically, in a federated setting where the pooled analysis is not feasible, our method provides a privacy-preserving solution that only requires a small amount of aggregated data to be shared across sites. Our method requires limited numbers of communications across sites and thus can be applied to both the automated and manual federated settings. In a centralized setting where the pooled analysis is allowed, our method can greatly reduce the computation time and memory cost. In all settings, our method can achieve nearly identical results as the gold-standard pooled analysis estimator, allows flexible specification of random and fixed effects in models, and can account for heterogeneity in the distribution of data across sites. We demonstrate the utility of Fed-GLMM through a real-world EHR data analysis that assesses characteristics associated with virtual versus in-person care utilization during the COVID-19 pandemic, using data from 8 healthcare facilities in the New England area. While the development of Fed-GLMM is motivated by EHR data analysis, the method can be used to address correlated structure in many types of real-world datasets, including administrative claims, genetic and clinical trial datasets, thus helping inform a wide range of clinical and scientific questions.

## 2 Results

### 2.1 GLMM Accounting for Site-Level Heterogeneity

Fed-GLMM allows modeling correlated observations from multiple EHRs using the following generic GLMM:

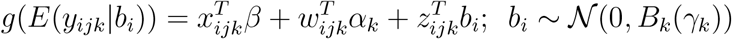

where *g*(.) denotes a link function, and *y*_*ijk*_ denotes the outcome variable of the *i*-th visit for the *j*-th patient at the *k*-th site. *x*_*ijk*_, *w*_*ijk*_ and *z*_*ijk*_ denote the corresponding covariates with common fixed effect *β*, site-specific fixed effect *α*_*k*_ and random effect *b*_*i*_, respectively. Note that *z*_*ijk*_ is a subset of the union of *x*_*ijk*_ and *w*_*ijk*_. The random effect *b*_*i*_ can be flexibly specified to account for different correlation structures. For example, we can include patient-level random effects to account for the correlation between visits of the same patient, and also physician-level random effects to account for the correlation between visits with the same physician.

To account for the heterogeneity across sites, we allow site-specific fixed effect *α*_*k*_ and site-specific variance-covariance structure *B*_*k*_, which is parameterized by *γ*_*k*_. In a homogenous setting, *α*_*k*_ and *B*_*k*_(*γ*_*k*_) can be set equal across sites. Compared with existing work where site-level heterogeneity is adjusted by introducing a random effect *b*_*k*_ [30–33], our method imposes no assumptions on the exchangeability of the site-level effects, which is more robust when the heterogeneity is large, and allows accurate estimation even when the number of sites is very small.

### 2.2 Fed-GLMM

The core concept upon which Fed-GLMM is built is to construct a quadratic surrogate function using summary statistics collected from each site to approximate the global likelihood function constructed from directly pooling all the data. Figure 1 provides an overview of the Fed-GLMM algorithm. We start with initialization for all the model parameters, denoted by 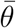. Since our model has both common parameters across sites and site-specific parameters, each site is required to fit its own GLMM in the initialization step. The initial values for the site-specific parameters are set to their local estimates (denoted by 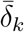 for the *k*-th site), while initial values for the common parameters are updated by a meta-analysis (denoted by 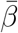). With more accurate initial values, we can achieve the same level of estimation accuracy with fewer communications across sites. In step 2, each site calculates and broadcasts summary statistics *s*_*k*_ and *H*_*k*_ involving less than *p*^2^ numbers (*p* is the number of parameters in the local model). These summary statistics are essentially derivatives of the local likelihood function. In step 3, the summary statistics obtained from step 2 are used to construct a quadratic surrogate function and obtain the parameter updates. When iterative communications are allowed, steps 2-3 can then be repeated to further update the parameter values. We present further details in the Methods section.

**Figure 1:**
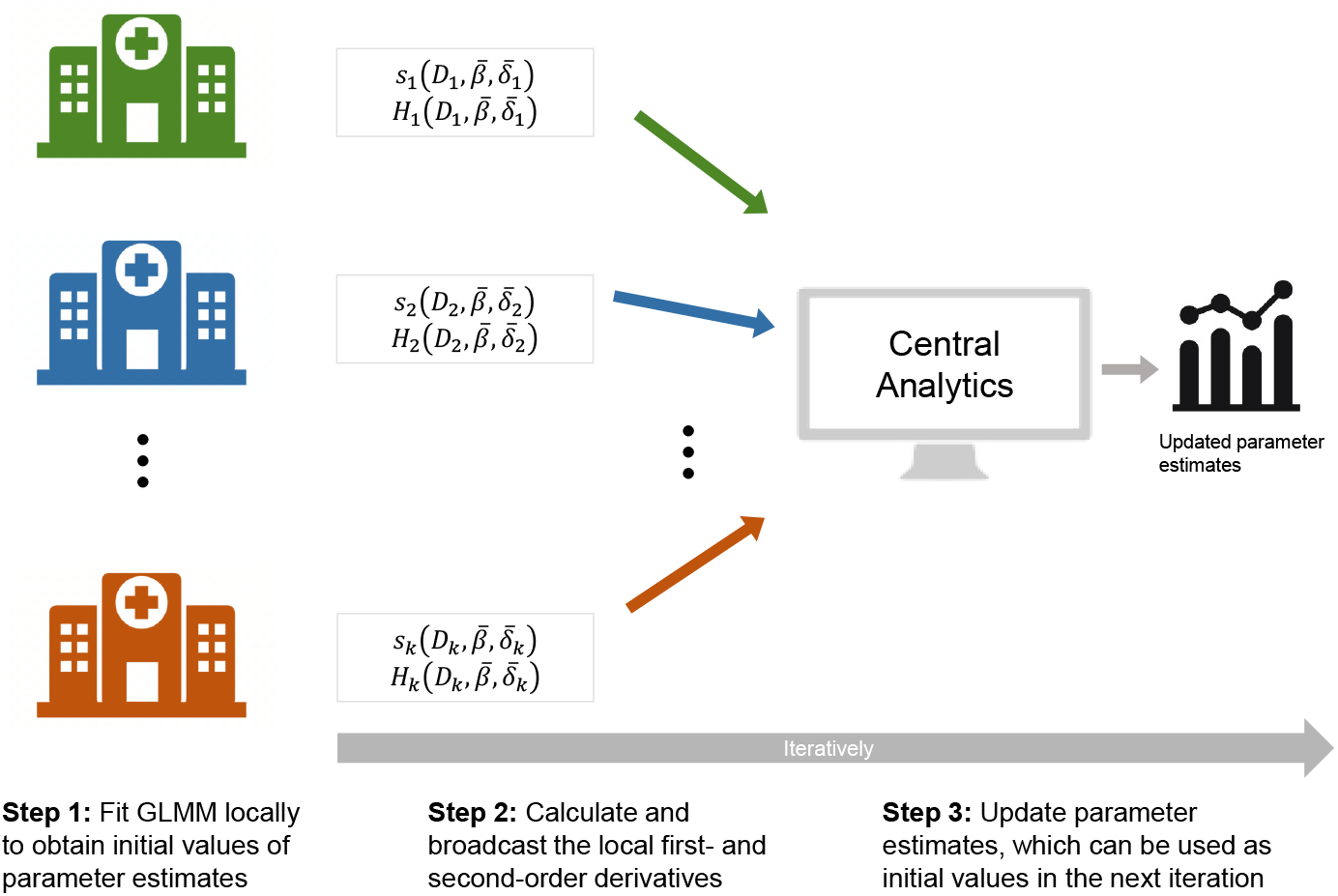
Schematic overview of Fed-GLMM. Fed-GLMM enables the joint implementation of GLMM for EHRs from multiple sites without sharing individual-level data. In step 2, each site calculates intermediate results s_k_, H_k_ locally, which are summary statistics evaluated at the initial values, and broadcasts them to the central analytics. For the k-th site, both s_k_ and H_k_ are functions of the local data D_k_, the common parameter value 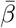, and the site-specific parameter value 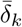. The local data D_k_ is composed of the local design matrix for the common fixed effect X_k_, the local design matrix for the site-specific fixed effect W_k_, and the local outcome vector y_k_. The site-specific parameter value 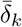 is composed of the values of site-specific fixed effect 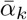 and site-specific variance parameter 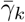. In step 3, the central analytics combines all the local intermediate results to construct a surrogate global likelihood function that provides updates for parameter estimates. Steps 2-3 can be iteratively performed to keep updating parameter estimates.

### 2.3 Simulation Study

We use a simulation study to demonstrate the improved accuracy of Fed-GLMM compared to the standard meta-analysis, and the improved computation time compared to the gold-standard pooled analysis. We generated simulated data mimicking a setting composed of 10 EHR datasets each with 100 patients and 5 visit encounters per patient. For each encounter, a binary outcome, a binary exposure and three additional covariates are generated from a GLMM with the logit link function, a patient-level random intercept, and a site-specific slope for one of the covariate variables. The detailed parameter and model specifications are described in the Methods section. We compared the estimation accuracy of the meta-analysis and Fed-GLMM using the relative bias from the pooled analysis estimator (referred to as the *relative bias* hereafter). We also compared the computational efficiency of Fed-GLMM to the meta-analysis and the pooled analysis.

#### Fed-GLMM demonstrated improved accuracy compared to meta-analysis in a federated setting

As shown in Figure 2 (upper left panel), the relative bias of the meta-analysis estimator for the exposure coefficient was more severe with rare outcomes or exposures. In contrast, Fed-GLMM converged to the values nearly identical to the pooled analysis estimates within 5 iterations in all prevalence settings. Additionally, in most non-rare event settings, Fed-GLMM achieved considerable improvement over the meta-analysis within 1-2 iterations. Compared with the meta-analysis, Fed-GLMM also demonstrated small variability in bias from the pooled analysis estimates across the simulation replicates, as shown in Supplementary Figure 1.

**Figure 2:**
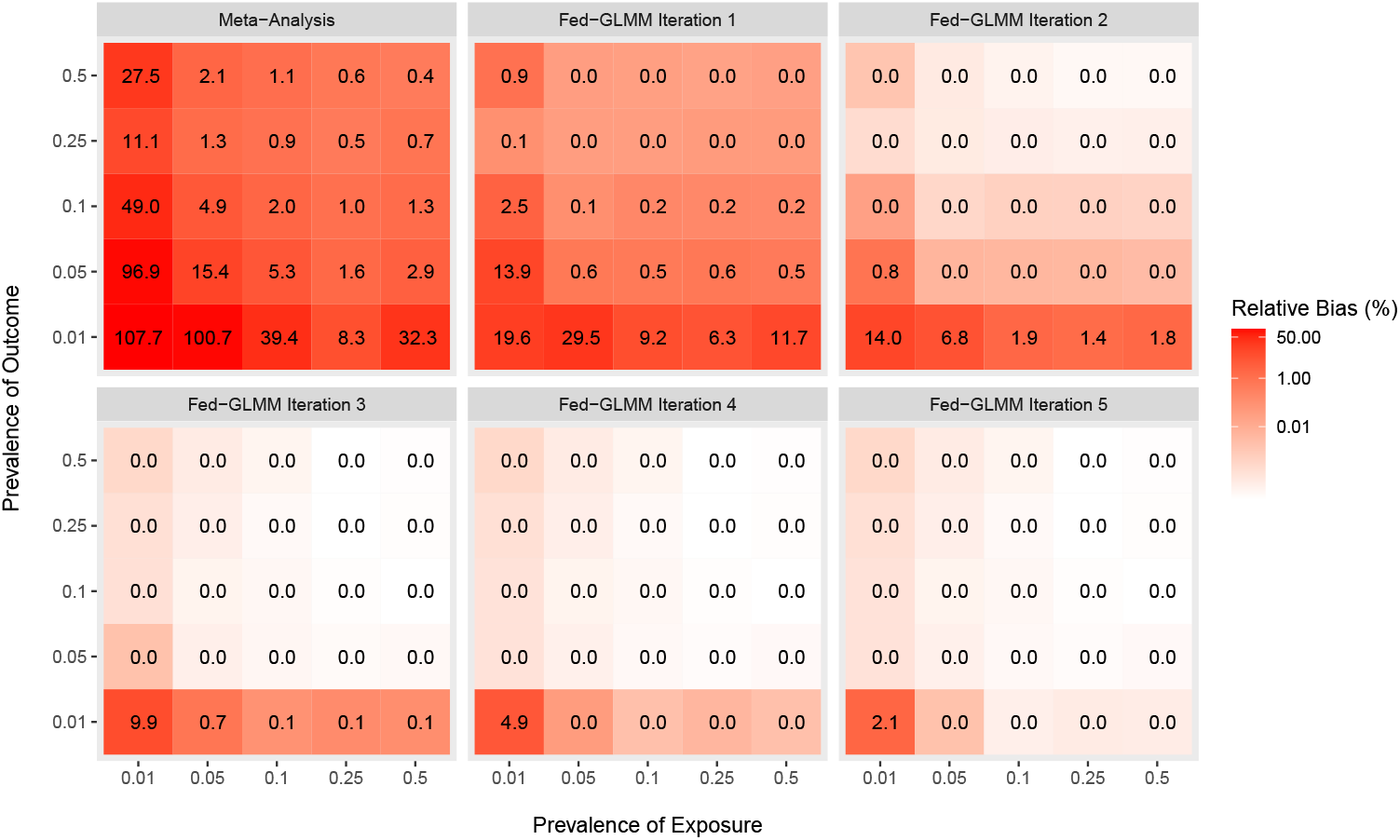
Accuracy of Fed-GLMM and meta-analysis estimates relative to gold-standard pooled analysis. We compared Fed-GLMM with the meta-analysis in their accuracy by calculating the median absolute relative difference from the gold-standard pooled analysis for the exposure coefficient estimate. The underlying model has a binary outcome, a binary exposure, three more covariates with 10 site-specific fixed effect coefficients for the normally distributed covariate and a patient-level random intercept. We considered 25 combinations of outcome and exposure prevalence to assess the model accuracy with 100 simulation replicates per combination. Fed-GLMM demonstrated reduced relative bias after 1-2 iterations compared with the meta-analysis, which was severely biased in the presence of rare events.

#### Fed-GLMM demonstrated improved computational efficiency compared to the pooled analysis in a centralized setting

We divided a pooled dataset into a different number of subsets and Fed-GLMM was applied using multiple computing nodes in parallel. Figure 3 shows that Fed-GLMM spent less than 5% of the computation time required by the pooled analysis when the number of computing nodes exceeds 20, and the time can be further reduced with more computing nodes. The meta-analysis can also provide a similar time reduction effect through parallel computing. However, with more computing nodes, the meta-analysis resulted in increasing relative bias, while Fed-GLMM retained its accuracy relative to the pooled analysis.

**Figure 3:**
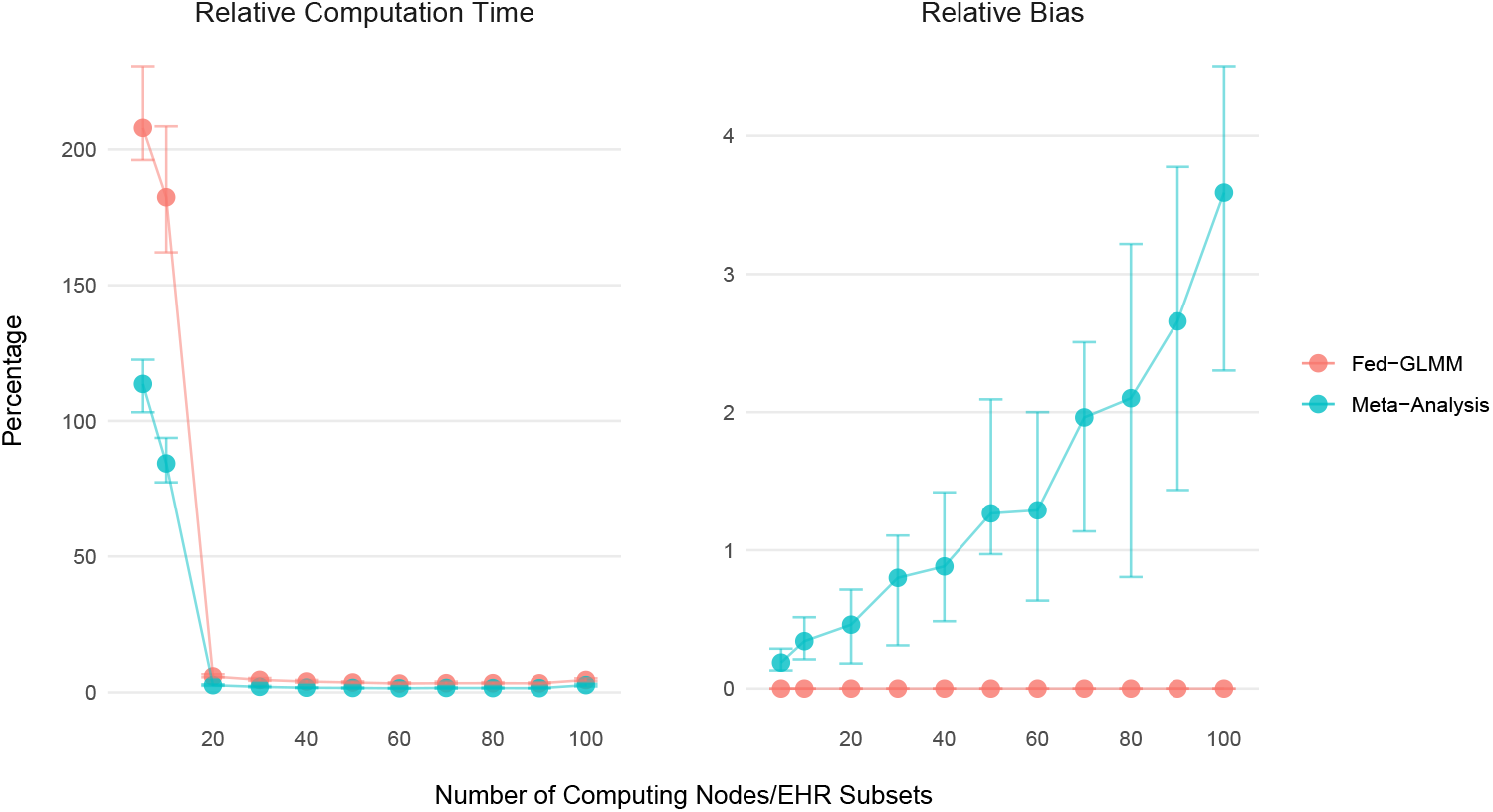
Comparison of computation time and estimate accuracy for Fed-GLMM and meta-analysis relative to gold-standard pooled analysis with increasing computing nodes/EHR subsets. We compared Fed-GLMM with the meta-analysis using the ratio (in percentage) of computation time over the pooled analysis. For each simulation replicate, we generated one single centralized EHR. The underlying model has a binary outcome, a binary exposure, three more covariates and a patient-level random intercept. We considered dividing the centralized EHR data into varying numbers of subsets to be computed in parallel. Both Fed-GLMM and the meta-analysis spent less than 5% of the computation time required by the pooled analysis with the number of computing nodes greater than 20. However, the meta-analysis had increased relative bias for the exposure coefficient when the number of subsets increased, while Fed-GLMM retained its accuracy relative to the pooled analysis. The points and bars represent median and interquartile range of computation time and relative bias in percentage respectively.

### 2.4 An Application of Fed-GLMM to Real-World EHR data: Evaluate Visit Characteristics Associated with Virtual Care Utilization

We applied Fed-GLMM to assess visit characteristics associated with virtual versus in-person care utilization during the COVID-19 pandemic. The outcome was defined as whether a care encounter or visit was conducted virtually versus in-person. The covariates of the model included variables measuring patient demographics, insurance status, English proficiency, digital literacy, visit type, and a temporal indicator for the “social normalization” during the COVID-19 pandemic. As care encounters are clustered by patients and physicians, we included both patient-level and physician-level random intercepts to account for the correlated observations. The detailed model specifications are described in the Methods section.

We identified all outpatient encounters over a one-year period between 10/1/2020 and 9/30/2021 from 8 acute care hospital facilities in the New England area. Combining data from 8 facilities, a total of around 3 million outpatient records are available. The facility with the highest visit volume has 1,194,009 records, making it computationally difficult to fit a GLMM even within one site.

#### Federated Setting

To demonstrate the use case of Fed-GLMM in a federated setting, we applied it to EHRs from all 8 facilities to fit the GLMM with facility-specific fixed intercepts that account for heterogeneity across facilities. EHRs from the two large facilities both with over 900,000 records were each split into 10 subsets to improve computation time, while other smaller EHRs were not split. Different from the simulation study where we had only the patient-level random effects, physician-level random effects were also included, which added difficulties in terms of dividing data into subsets. If patients were nested within physicians, we could simply split the records by physicians. However, since multiple visits of the same patient can be associated with different physicians, a random division based on physicians is not the optimal splitting strategy as observations of the same patient would be split into different subsets. We proposed a clustering-based splitting method that splits physicians based on how many shared patients they have. Two physicians are more likely to be assigned to the same subsets if they share more patients. In this way, we can best preserve the correlation structure of the data, and therefore achieve better accuracy. A numerical evaluation has shown that the clustering-based splitting strategy has higher estimation accuracy than the random splitting strategy. More details of the evaluation are shown in Supplementary Figure 2.

#### Centralized Setting

We demonstrate the computation time benefit of Fed-GLMM compared to the pooled analysis in a centralized setting. We fit the same GLMM using the EHR data from the facility with the highest visit volume. With Fed-GLMM, the EHR was split into 10 smaller subsets to be computed in parallel. Fitting Fed-GLMM to the EHR data of the single facility (n = 1,194,009) using the clustering-based splitting strategy, we observed convergence at the 4th iteration. The entire process took 5,026 seconds, while the analysis would be otherwise infeasible if all data were fit in a single GLMM process. Our analysis was performed with R 4.0.2 on a Linux cluster with up to 512GB RAM per node at 1600MHz.

The results of EHR modeling for the federated (all 8 facilities) and centralized (the facility with the highest visit volume only) settings are summarized in Figure 4. In both settings, the characteristics associated with lower odds of conducting a virtual visit (i.e., greater odds of in-person visit) includes increasing age, Hispanic, non-Hispanic black, non-Hispanic Asian or other non-Hispanic relative to non-Hispanic white race/ethnicity, limited English proficiency, inactive patient portal (as a proxy for lower-level of digital literacy). Compared with primary care visits, behavioral health and specialty visits were more likely to be conducted virtually. Visits of female patients, as well as visits billed to Medicaid were also more likely to be conducted virtually.

**Figure 4:**
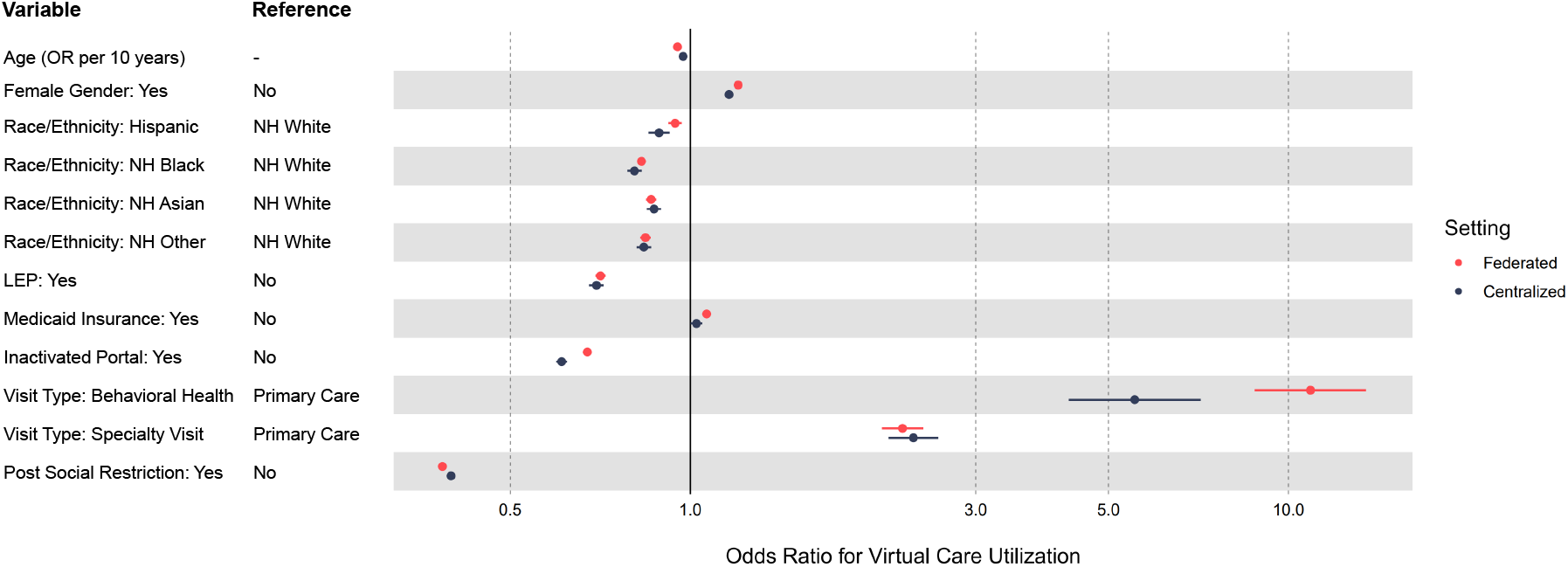
Adjusted odds ratios of virtual visit vs. in-person visit of patient and visit characteristics. Using the forest plot, we visualized the adjusted odds ratios obtained through Fed-GLMM for both all facilities (federated setting to demonstrate privacy preservation) and the single facility (centralized setting to demonstrate computation improvement). The points and bars represent the point estimates and 95% confidence intervals, respectively. **Abbreviations:** OR - Odds Ratio; NH - Non-Hispanic; LEP - Limited English Proficiency

## 3 Discussion

In light of the increasing need for multicenter collaborative research utilizing EHR data, and the potential challenges in data sharing and large-scale computation, we proposed the Fed-GLMM algorithm to model correlated EHR data that allows privacy-preserving integration of datasets from multiple healthcare systems. Our method also enables fitting GLMM with much less computation time and memory cost in both federated and centralized networks, and thus can also be applied to EHR from a single site. Our simulation study has demonstrated that Fed-GLMM achieves nearly identical results to the pooled analysis with reduced computation time over a broad spectrum of settings. Our real-world data analysis demonstrated the feasibility of applying Fed-GLMM to single-site and multicenter EHR-based studies to fit a model with millions of observations.

Compared to existing work, the most important contribution of Fed-GLMM is that it allows the modeling of longitudinal and correlated data within each institution and can accommodate all GLMM specifications, including crossed or nested random effects. However, when performing Fed-GLMM to improve computational efficiency through splitting large-scale data, one needs to be mindful of the data splitting strategy to generate accurate results for models with crossed or nested random effects. For nested random effects, splitting the data by the highest-level factors will allow Fed-GLMM estimates to converge to the gold-standard pooled analysis results. For crossed random effects, one should split the data such that the correlated observations are allocated to the same subsets as much as possible as shown in Supplementary Figure 2. This makes the Fed-GLMM estimates close (though not identical) to the pooled analysis estimates.

As demonstrated in our simulation and real-world data analyses, iterative communication among the central analytics and individual sites is not required. In most cases, only one round of parameter updating provides negligible bias. Thus, in federated research networks that rely on manual data transferring, our method with one round of iteration is preferred to reduce the communication cost. However, when multiple rounds of communication are feasible, with an increasing number of iterations, our method will eventually converge to the pooled analysis. When studying rare conditions, extra iterations help correct the bias, so a balance needs to be reached between the communication cost and estimation accuracy. In addition, the sharing of first- and second-order derivatives is common among federated algorithms but may still entail a risk of identifiability for small datasets with rare events. Nevertheless, this risk is limited in that the transmission of summary-level statistics is typically regulated and protected by the data-sharing protocols of collaborative research networks. Methods such as differential privacy and data encryption techniques can be combined with Fed-GLMM to improve privacy protection.

In the virtual care analysis, we found lower odds of virtual care use among Hispanic, non-Hispanic black, non-Hispanic Asian or other non-Hispanic relative to non-Hispanic white patients but higher odds among patients with Medicaid insurance. These results are in contrast to a previous study investigating the same health system [34], which used a generalized linear model without accounting for the patient- and physician-level correlations. The different findings may be explained by distinct study periods and different levels of virtual care adoption over time, or may be explained by the different methodological approach we employed here. By including random effects, our analysis was better able to address correlations among visits within patients and physicians. While we have demonstrated Fed-GLMM for analyzing EHR data to assess virtual care utilization, the algorithm can be used in other types of datasets with correlated observations to investigate a variety of biomedical and epidemiological research questions.

## 4 Methods

### 4.1 Fed-GLMM Algorithm

Suppose we want to use Fed-GLMM to integrate data from *K* EHRs stored at *K* different sites. At each site, we fit the following model:

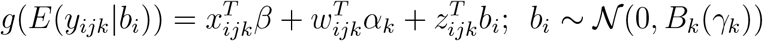

where *β* denotes the fixed effects shared across sites, *α*_*k*_ denotes the fixed effects specific to the *k*-th site, and *B*_*k*_ denotes the variance-covariance matrix of the random effect *b*_*i*_ and is parameterized by *γ*_*k*_.

Let *I*_*k*_ denote the index set indicating patients in the *k*-th site. Suppose the *j*-th patient has *n*_*j*_ observations. The log-likelihood function constructed by data from the *k*-th site can then be written as

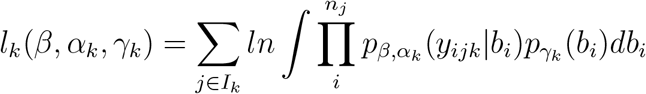

Since the integral above does not have a closed-form solution, the log likelihood is often approximated by methods such as the penalized quasi-likelihood, Laplace’s method or Gaussian quadrature [27, 28]. Fed-GLMM applies to any of these integral approximation methods, and here we use Laplace’s method as an example which approximates the log likelihood by

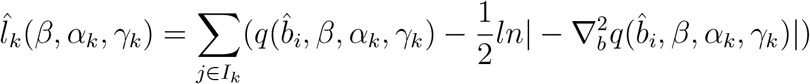

where 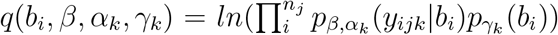 with *b*_*i*_ evaluated at 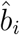 such that the corresponding first-order derivative 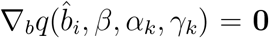, and 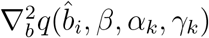 denotes the second-order derivative.

We denote the parameters specific to the *k*-th EHR as *δ*_*k*_ = (*α*_*k*_, *γ*_*k*_), and denote the entire set of parameters as *θ* = (*β, δ*_1_, *δ*_2_, … , *δ*_*K*_)^*T*^ . The combined log-likelihood function encompassing all *K* EHRs can be written as

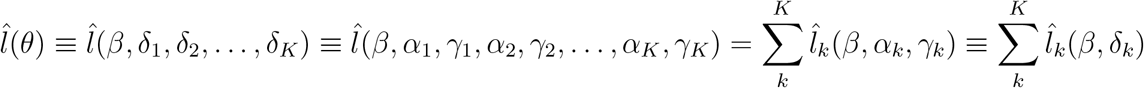

We then propose the following quadratic surrogate function that approximates the global function 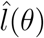 at an initial value 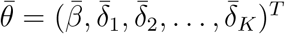:

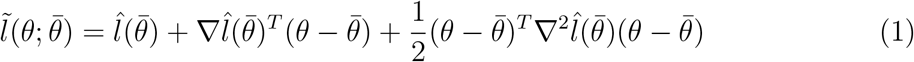

To obtain 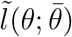, site *k* needs to share

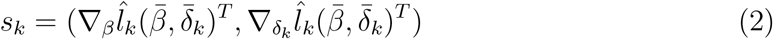

and

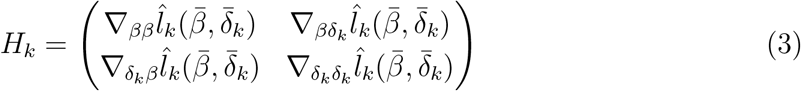

which are summary statistics that can be calculated using the local EHR given an initial value 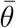. After getting all the summary statistics, the parameter estimates can be updated through 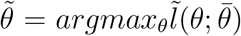. The above procedure can be repeated *T* times when iterative communications are allowed or until a convergence criterion *d* is reached. We denote the resulting value as *θ*_*Fed*_. The Fed-GLMM algorithm is summarized as Algorithm 1.

#### Algorithm 1: Fed-GLMM

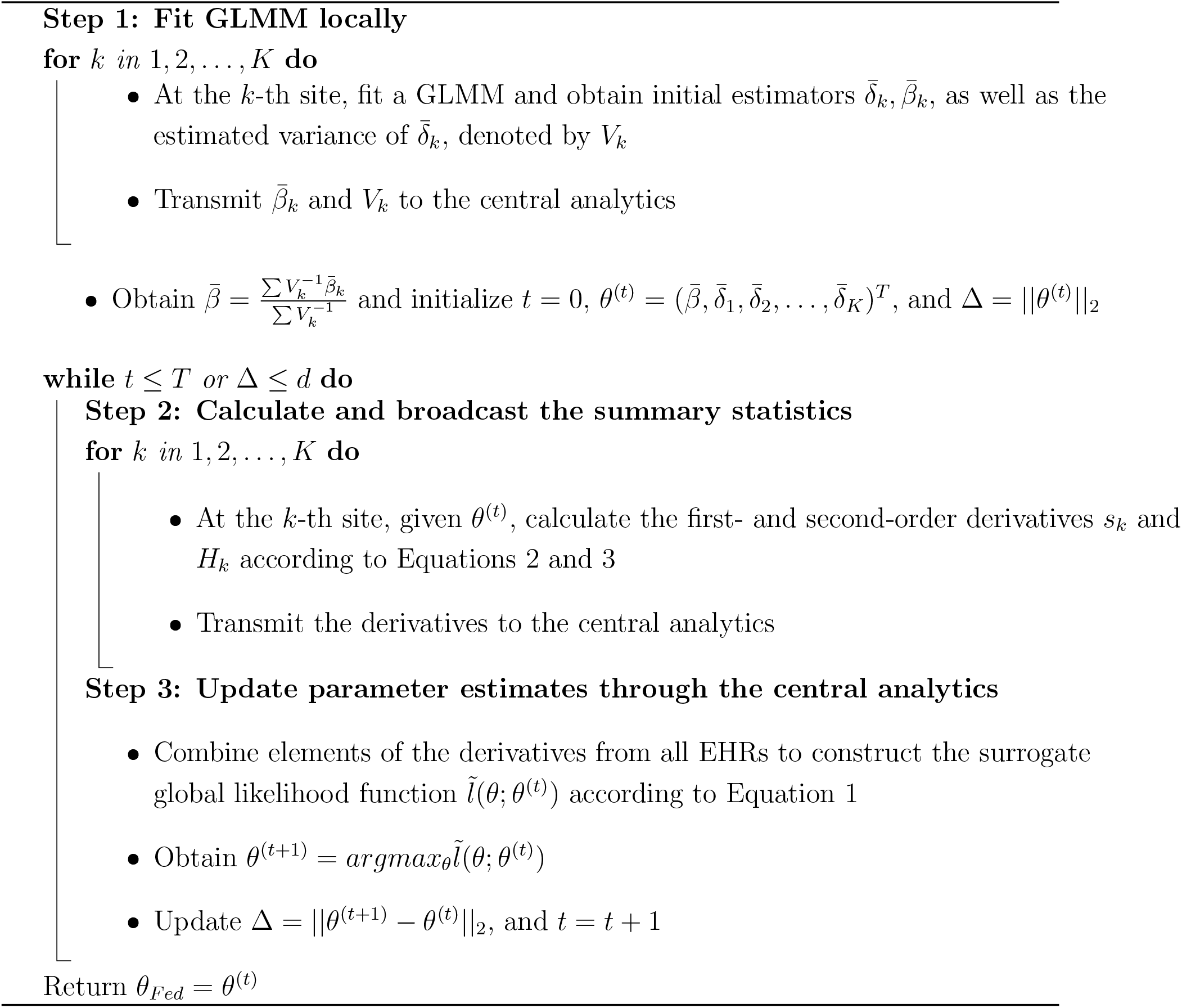

The algorithm also applies to the centralized setting in which site-specific parameters are not involved and our model of interest becomes

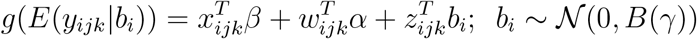

where the entire set of parameters is redefined as *θ* = (*β, α, γ*)^*T*^ .

### 4.2 Variance Estimation

The variance of the Fed-GLMM estimator can be calculated directly using the Hessian of the surrogate global likelihood evaluated at *θ*_*Fed*_, denoted as *H*_*Fed*_. The variance-covariance estimator for *θ*_*Fed*_ can then be obtained through 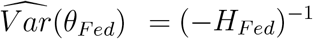.

### 4.3 Simulation: Evaluate the Accuracy of Fed-GLMM

We considered a GLMM with a binary outcome, a binary exposure and three additional covariates (one binary and two continuous variables that follow standard normal and uniform distributions respectively). The model also included a patient-level random intercept, which can be expressed as

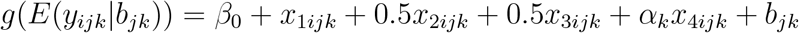

where *x*_1*ijk*_ ∼ *Bernoulli*(*p*_*x*_), *x*_2*ijk*_ ∼ *Uniform*(0, 1), *x*_3*ijk*_ ∼ *Bernoulli*(0.5), and *x*_4*ijk*_ ∼ 𝒩 (0, 1).

We randomly assigned *k* distinct values ranging from 0 to 1 for the *K* sites as the site-level fixed effect *α*_*k*_. To study the impact of the prevalence of binary exposure on the model performance, we let *p*_*x*_ vary from 0.01 to 0.5. By choosing different values of *β*_0_, we were also able to allow the prevalence of the binary outcome to vary from 0.01 to 0.5.

In a single simulation replicate, we simulated 10 EHR datasets, each with 100 patients and 5 encounters per patient (500 encounters in total). We performed the pooled analysis, the meta-analysis and Fed-GLMM respectively. We evaluated the accuracy of Fed-GLMM vs. meta-analysis using the absolute relative bias from the pooled analysis for estimating the coefficient of the binary exposure *x*_1_, calculated as the following:

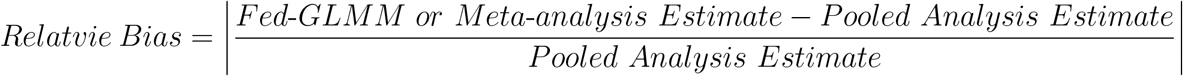

The simulation results are summarized in Figure 2 and described in the Results section.

### 4.4 Simulation: Evaluate the Computational Efficiency of Fed-GLMM

We considered a homogeneous setting with the same GLMM specifications and variable distributions as in the previous simulation. In a single simulation replicate, we generated a single dataset with 5,000 patients and 5 encounters per patient (25,000 encounters in total). The prevalence of the binary exposure was set to 0.05 and the prevalence of binary outcome was set to 0.25. We applied Fed-GLMM by randomly splitting the dataset into subsets. We investigated the computation time with the number of subsets ranging from 5 to 100. We generated 50 simulation replicates for each setting, and for each simulation replicate, we also performed the pooled analysis and the meta-analysis. We evaluated the computational efficiency of Fed-GLMM and the meta-analysis as the ratio of their computation time to that of the pooled analysis, calculated as the following:

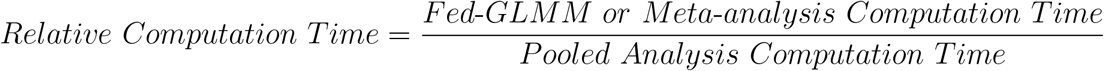

The simulation results are summarized in Figure 3 and described in the Results section.

### 4.5 Real-World Data Analysis

The Virtual Care dataset was curated from the data warehouse of a large New England healthcare system. For a demonstration of Fed-GLMM, we included all 3,165,913 ambulatory visits with physicians conducted at 8 acute care hospital facilities during a one-year period from 10/1/2020 through 9/30/2021. We extracted patient characteristics and demographics, physician primary specialty, and whether the visit was conducted in person or virtually through associated modifier codes. We performed a complete-case analysis where all observations with missing values (6.8% or 215,329 visits) were excluded from the final analytical sample. This EHR study was approved by the Mass General Brigham Institutional Review Board as a medical record review that did not require patient consent.

We considered a GLMM with a binary outcome indicating whether a care encounter was conducted virtually (coded as 1) or in-person (coded as 0). The covariates in the model included patient age, gender, race/ethnicity (Hispanic, non-Hispanic white, non-Hispanic black, non-Hispanic Asian and other non-Hispanic race/ethnicity), English proficiency (whether the patient indicated English as the preferred language), the digital patient portal status (whether the portal was activated or not for the care encounter as a proxy for “digital literacy”), insurance status (whether the visit was billed to the Medicaid as a proxy for social determinants of health), visit type (whether the visit was completed in a primary care, behavioral health or specialty department), and an indicator for whether the care happened on or after 5/29/2021 – the ending of social restriction in Massachusetts (to approximate the beginning of “social normalization” during the COVID-19 pandemic in the New England area). All covariates were entered in the model linearly. We included a physician-level random intercept and a patient-level random intercept to account for the correlations across encounters at different levels.

When applied to the centralized large EHR from a single facility (n=1,194,009) which was split into 10 subsets, Fed-GLMM fit the model above for each subset separately. With two crossed random effect terms in the model, Fed-GLMM provides accurate results only when the data can be appropriately split such that the correlated observations are allocated to the same data subsets as much as possible. One way to achieve this is to cluster physicians who shared patients together and then split data by physician clusters. We examined the crossing structure of patients and physicians through physicians’ patient-sharing network. While existing network community detection algorithms can be handy to cluster physicians, desired cluster numbers and uniform cluster sizes are not always achievable for densely connected networks. Therefore, we proposed an algorithm, summarized as Algorithm 2, to implement the clustering-based splitting strategy.

#### Algorithm 2: Clustering-based Data Splitting Strategy for Crossed Patient and Physician Random Effects

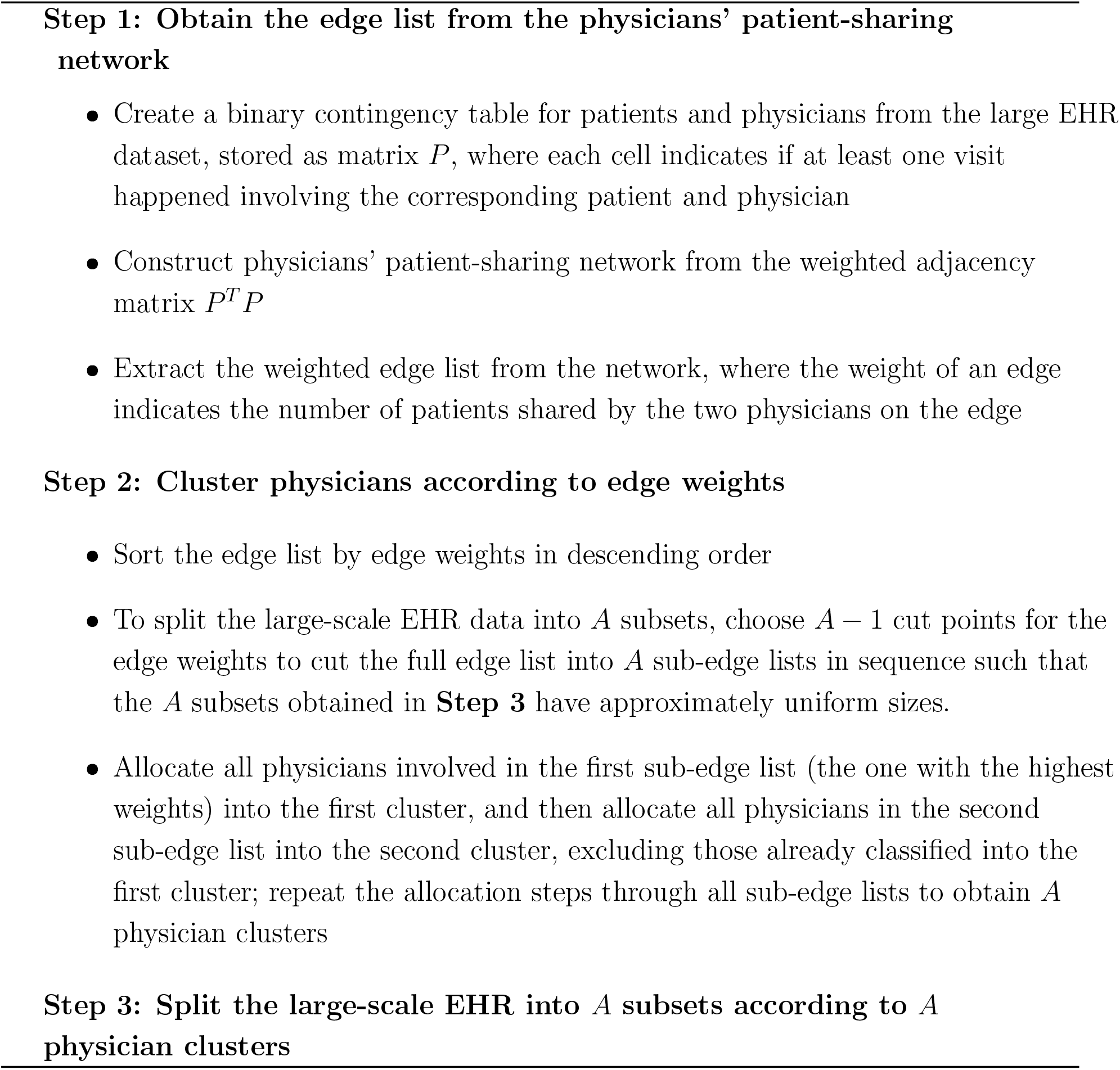

When applying Fed-GLMM to EHRs from all 8 facilities, we included facility-specific intercepts (fixed effects) to account for the potential heterogeneity in the prevalence of the virtual visits across facilities. EHRs from the two large facilities both with over 900,000 records were each split into 10 subsets using the clustering-based data splitting approach to improve computation time, while other smaller EHRs were not split.

## Data Availability

Because the dataset contains protected health information, we are unable to share it in full. Upon reasonable request, we may be able to share a deidentified version with appropriate approvals from our institution and IRB.

