## Supplementary Materials for "Fed-GLMM: A Privacy-Preserving and Computation-Efficient Federated Algorithm for Generalized Linear Mixed Models to Analyze Correlated Electronic Health Records Data"

**Supplementary Figure 1: Distribution of Fed-GLMM and meta-analysis estimates relative to gold-standard pooled analysis across simulation replicates.**

**Supplementary Figure 2: Comparison of Fed-GLMM accuracy relative to pooled analysis between different data splitting strategies with repeated small samples from a single large-scale EHR.**

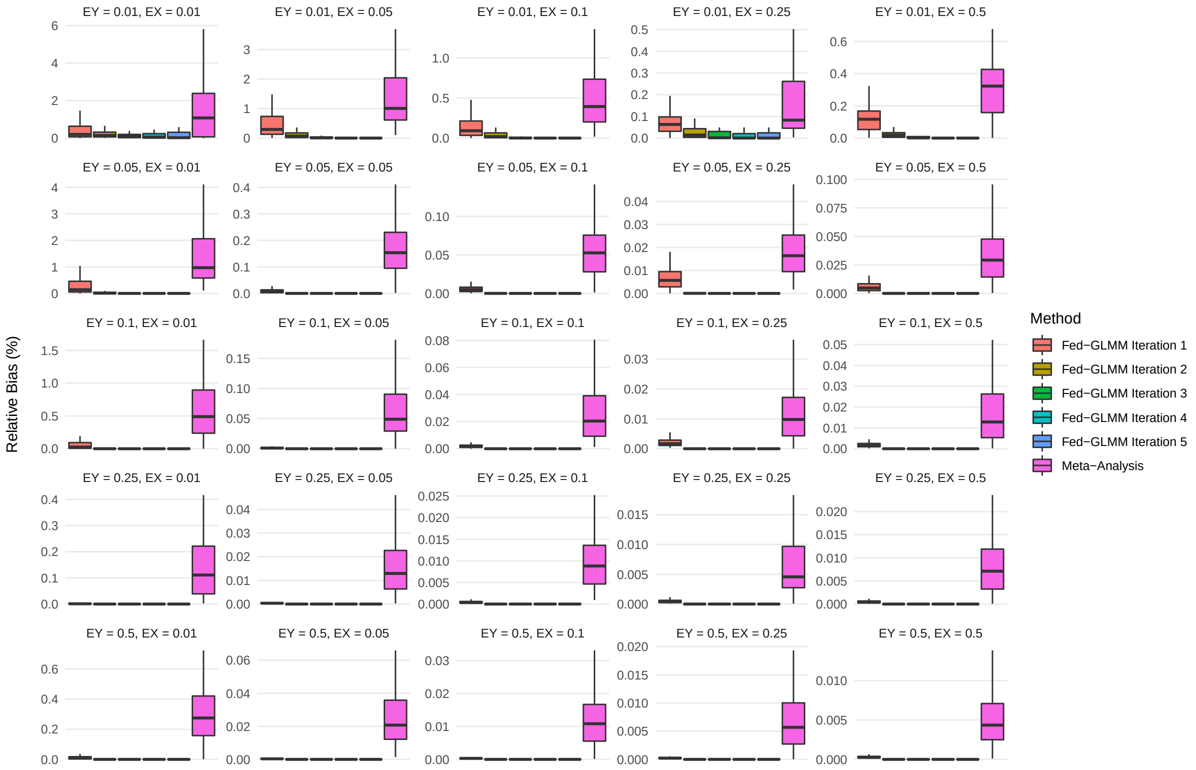


**Supplementary Figure 1: Distribution of Fed-GLMM and meta-analysis estimates relative to gold-standard pooled analysis across simulation replicates.** *We compared Fed-GLMM with the standard meta-analysis in accuracy by calculating the absolute relative difference from the gold-standard pooled analysis for the exposure coefficient estimtate. The underlying model has a binary outcome, a binary exposure, three more covariates with 10 site-specific fixed effect coefficients for the normally distributed covariate and a patient-level random intercept. We considered 25 combinations of outcome and exposure prevalence to assess the model accuracy with 100 simulation replicates per combination. Fed-GLMM achieved almost identical results as the pooled analysis for all simulation replicates after 1-2 iterations, while the meta-analysis demonstrated greater bias and variance relative to the pooled analysis across all simulation replicates and prevalence settings.* ***Abbreviations:*** *EY - Prevalence of Outcome; EX - Prevalence of Exposure*

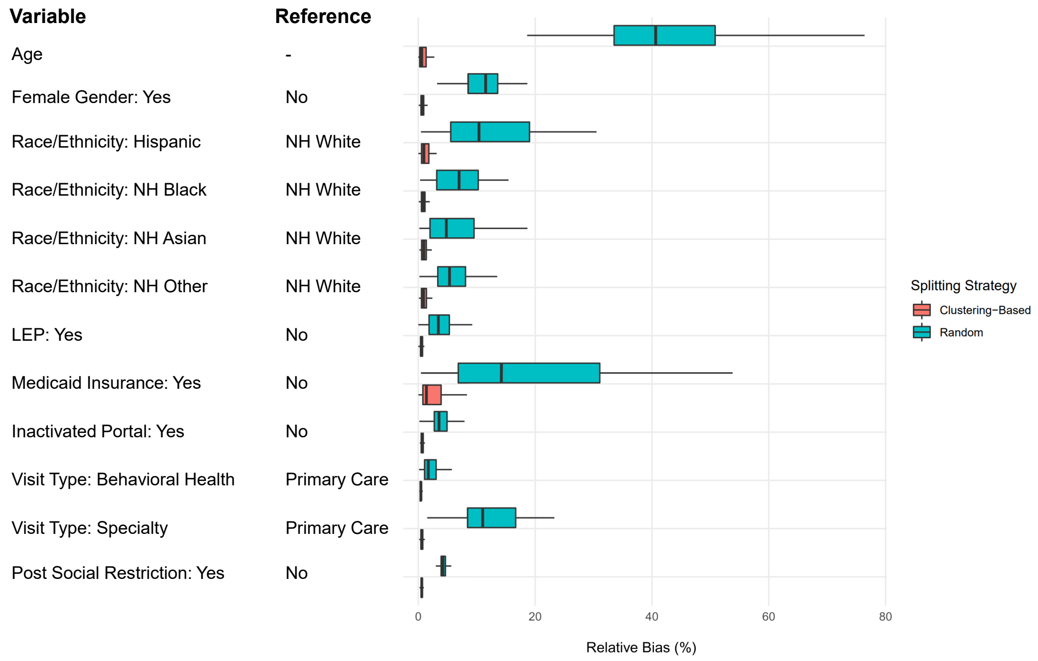


**Supplementary Figure 2: Comparison of Fed-GLMM accuracy relative to pooled analysis between different data splitting strategies with repeated small samples from a single large-scale EHR.** *We used small sub-datasets (n=100,000 for each randomly extracted replicate) of the EHR from a single facility to demonstrate the accuracy of Fed-GLMM with different data splitting strategies. Two splitting strategies were attempted and compared: random splitting and our proposed clustering-based splitting, which was designed to mitigate the impact of the separation of correlated observations into different subsets in the presence of crossed random effects. Each replicate was split into 5 subsets by both strategies. The bias was calculated as the difference between the corresponding Fed-GLMM estimates and those given by the pooled analysis in absolute percentage. For all coefficients of interests, clustering-based splitting resulted in negligible bias compared with the random splitting strategy.* ***Abbreviations:*** *NH - Non-Hispanic; LEP - Limited English Proficiency*
